# Use of traditional medicine in treatment of migraine during coronavirus disease 2019 (COVID-19) pandemic: an online survey

**DOI:** 10.1101/2021.05.25.21257676

**Authors:** Ismail Ibrahim Ismail, Jasem Y. Al-Hashel

## Abstract

Since the declaration of coronavirus disease 2019 (COVID-19) pandemic, patients with migraine were specifically vulnerable to worsening of their headaches. Traditional medicine (TM) has been used to treat headache disorders for centuries, especially during times of a healthcare emergencies, similar to the current pandemic. We aimed to assess the use of TM in treatment of migraine amid COVID-19 pandemic in Kuwait, using an online, self-administered questionnaire. A total of 1018 patients completed the survey. TM was used by 39.9% respondents. The greatest users of TM were those with older age (p =0.04), and longer disease duration (p =0.005). TM users were found to be more compliant to treatments than non-TM users (p<0.02). However, they reported significantly less communication with their physicians (p <0.001) during the pandemic. This study showed higher rates of TM use, as a way of self-treatment, among patients with migraine during COVID-19 pandemic. Neurologists should prepare their patients with “rescue” strategies for headache management, and new means of communication, to face these “*new normal*” challenges.

## Introduction

Since the emergence of coronavirus disease 2019 (COVID-19) pandemic, patients with migraine were specifically vulnerable to worsening of their headaches, in addition to inadequate quality of medical care and overall psychosocial stressors (1,2). Traditional medicine (TM) has been used for centuries to treat headache disorders, and around 2/3 of migraine patients reported seeking TM instead of conventional medicine in literature (3,4).

## Main text

We conducted an internet-based, cross-sectional study, to assess the use of TM in treatment of migraine amid COVID-19 pandemic. A self-administered questionnaire was distributed to patients with migraine from the headache clinic in the largest neurology center in Kuwait, and was also posted on several medical social-media accounts. The study was conducted between July 15 and July 30, 2020.

A total of 1018 patients completed the survey. TM was used by 406 (39.9%) respondents; 353 (86.9%) females, with mean age of 34 ± 9.5 years, and mean disease-duration of 9.8 ± 8.3 years.

The reported methods during the pandemic were; head massage by 533 (52.3%) respondents, head banding by 488 (47.9%), herbal drinks (hibiscus, tamarind, ginger, chamomile, cinnamomum cassia, coriander seed, valerian, nigella sativa “black cumin”, saussurea costus, coffee, tea) by 467 (45.8%), essential oil aromatherapy (lavender oil, peppermint oil, rosemary oil, eucalyptus oil, anise oil) and “Migraine Sticks” by 379 (37.2%), local application of (Henna, Camphor oil cream (Vicks), Jujube/Sidr) by 267 (26.2%), ice packs by 234 (22.9%), supplements (magnesium, feverfew, butterbur, vitamin B2, vitamin C, vitamin D3, CoQ10) by 227 (22.3%), dietary changes (healthy diets, avoiding triggers, and eating food rich in magnesium as bananas, pumpkin seeds, almonds) by 201 (19.7%), good hydration by 181 (17.7%), relaxing techniques by 177 (17.3%), sleep hygiene by 156 (15.3%), aerobic exercises by 135 (13.2%), “*Sabkha*” (mixture of herbs that is heated and applied on top of the head for few days) by 111 (10.9%), hot compresses by 83 (8.1%), Ayurvedic medicine and Homeopathy by 51 (5.0%), blood cupping (*Hijama*) by 12 (1.1%), “*Farry*” treatment (folks’ belief of a gap in the skull that is treated by head squeezing by traditional healers) by 5 (0.5%), yoga by 4 (0.4%), and acupuncture by 2 (0.2%) respondents.

Participants who reported higher migraine frequency and migraine severity were found to use TM the most; (44.5% vs 33.1%, p <0.001) and (43.5% vs 33.4%, p=0.002), respectively.

The greatest users of TM were those with older age (44 ± 7.5 vs 32 ± 8.5, p =0.04), and longer disease duration (12.6 ± 7.3 vs 8.8 ± 8.6, p =0.005). Interestingly, TM users were more compliant to their treatments than non-TM users (52.5% vs 43.1%, p<0.02). However, they reported significantly less communication with their physicians during the pandemic (33.9% vs 55.3%, p <0.001).

This survey provides interesting insights on how migraine patients attempted self-treatment with TM during the pandemic, as a result of cancellation of face-to-face visits and procedural treatments. In a recent study (5), 30.1% of migraine patients used TM while being treated by a neurologist. In our survey, the percentage increased to 40 %, despite the inability of most patients to undergo traditional procedures during lockdown, and for fear of infection.

## Conclusion

Worsening of migraine, in addition to the overall negative impact of COVID-19 pandemic, has forced the patients to self-treat themselves by using TM, despite being compliant to their conventional treatments. Neurologists should prepare their patients with “rescue” strategies for headache management, and new means of communication, to face these “*new normal*” challenges.

## Data Availability

All data are available with the corresponding author upon request

## Abbreviations

COVID-19: Coronavirus disease 2019
TM: Traditional medicine

## Declarations

## Ethics approval and consent to participate

The study was approved by the Scientific Research Committee of Department of Neurology, Ibn Sina hospital, Kuwait, the dedicated ethics oversight body, in July 2020. Answering the survey was considered an implied consent to participate in the study, as approved by the Scientific Research Committee of the department, based on the fact that completing the survey was voluntary, and all answers were confidential.

## Consent for publication

Informed consent to publish this information was obtained from study participant.

## Availability of data and materials

The data sets supporting the conclusion of this article are included within the article.

## Authors’ contributions

III and JYA had the original idea for the study, and planned the overall design. III wrote the initial draft, was responsible for data acquisition and analysis of the results, in addition to preparation of the final version of manuscript, making amendments at the review and post-review stage, and he is the first and correspondent author. JYA critically reviewed the initial manuscript. The authors have read, revised and approved the final manuscript.

## Acknowledgements

Not applicable.

## Authors’ information

III-MD, Neurology specialist

JYA-MD, Professor of neurology

